# Sociodemographic predictors of outcomes in COVID-19: examining the impact of ethnic disparities in Northern Nevada

**DOI:** 10.1101/2020.05.24.20112094

**Authors:** Daniel Antwi-Amoabeng, Bryce D. Beutler, Munadel Awad, Zahara Kanji, Sumaiya Mahboob, Jasmine Ghuman, Sri Harsha Boppana, Mohammad S. Sheikh, Mark B. Ulanja, Nageshwara Gullapalli

**Affiliations:** University of Nevada, Reno School of Medicine, Reno, NV

## Abstract

**Background:** On March 11, 2020, the World Health Organization declared coronavirus disease-19 (COVID-19) a pandemic. Nearly five million individuals have since been diagnosed with this increasingly common and potentially lethal viral infection. Emerging evidence suggests a disproportionate burden of illness and death among minority communities. We aimed to evaluate the effect of ethnicity on outcomes among patients diagnosed with COVID-19 in Northern Nevada.

**Design:** Single-center, retrospective observational study

**Materials and methods:** The electronic health records of 172 patients diagnosed with COVID-19 were obtained from a 946-bed tertiary referral center serving Northern Nevada. Demographic and clinical characteristics were compared by ethnic group (Hispanic versus non-Hispanic). Logistic regression was used to determine predictors of mortality.

**Results:** Among 172 patients who were diagnosed with COVID-19 between March 12^th^ and May 8^th^, 2020, 87 (50.6%) identified as Hispanic and 81 (47.1%) as non-Hispanic. The mean age was 46.0 among Hispanics and 55.8 among non-Hispanics. Comorbidities linked to increased COVID-19-related mortality – hypertension, obesity, and chronic obstructive pulmonary disease – were more common among the non-Hispanic population. Hispanic individuals were significantly more likely to be uninsured and to live in low-income communities as compared to their non-Hispanic counterparts (27.6% versus 8.2% and 52.9% versus 30.6%, respectively). Hispanic patients were also less likely than non-Hispanics to have a primary care provider (42.5% versus 61.2%). However, mortality was significantly higher among the non-Hispanic population (15.3% versus 5.8%).

**Conclusion:** The COVID-19 pandemic has disproportionately affected Hispanic individuals in Northern Nevada, who account for only 25.7% of the population but over half of the confirmed cases. Hispanic individuals were younger and had fewer comorbidities than their non-Hispanic counterparts; consequently, despite considerable socioeconomic disadvantage, mortality was lower among the Hispanic population. The underlying causes of ethnic disparities in COVID-19 incidence remain to be established, but further investigation may lead to more effective community- and systems-based interventions.

## INTRODUCTION

The coronavirus disease-19 (COVID-19) pandemic has changed the face of healthcare in the United States. As of this writing, over 1.5 million Americans have been diagnosed and nearly 100,000 have died of this increasingly common viral infection. Emerging evidence suggests a disproportionate burden of illness and death among communities of color [1-3]. The underlying causes of ethnic disparities in the setting of COVID-19 remain to be established, but it has been postulated that social distancing represents a privilege of the dominant ethnic group; minority individuals are more likely to work essential jobs and live in multigenerational households as compared to their white counterparts [4,5]. In addition, distrust of the medical establishment is pervasive among minority communities, and public health messages may not resonate among individuals who have been affected by systemic racism [6].

Ethnic disparities in COVID-19 outcomes have been reported by other authors. However, there is a paucity of data pertaining specifically to the Hispanic population. In this study, we aimed to compare COVID-19 incidence and outcomes between the Hispanic and non-Hispanic populations of Northern Nevada.

## METHODS

### Source of data

We conducted a single-center, retrospective observational study using data extracted from the electronic health records of Renown Regional Medical Center – a 946-bed tertiary referral hospital serving most of Northern Nevada – and affiliated satellite hospitals. The study protocol was approved by the institutional review board of the University of Nevada, Reno School of Medicine.

### Study population

The study sample included all patients who had a positive nasopharyngeal reverse-transcriptase polymerase chain reaction test for severe acute respiratory syndrome coronavirus 2 (SARS-CoV-2) between March 12^th^ and May 8^th^, 2020 (N = 172).

### Variables

The primary outcome of interest was mortality. Secondary outcomes included the following: hospitalization, acute respiratory distress syndrome (ARDS), sepsis, shock, mechanical ventilation, hepatic injury, renal injury, arrhythmia, coagulopathy, and hemorrhage (as defined by the International Classification of Diseases, 10^th^ revision [ICD-10] coding). Additional patient-level data included age, sex, comorbidities, ethnicity (Hispanic or non-Hispanic), income class (low or middle/upper), insurance type (Medicare, Medicaid, private, uninsured, or workers’ compensation), and the presence or absence of a primary care provider.

### Statistical analyses

Categorical variables were described using proportion and were compared among ethnic grouping with Fisher’s exact test. Continuous variables were not normally distributed and were described as medians and interquartile ranges (IQRs). Sex, age categories, ethnic grouping, presence of diabetes, hypertension, obesity, chronic kidney disease, chronic obstructive pulmonary disease, and intensive care unit stay were used as predictors to fit a logistic regression model for the outcome of death. The Hosmer-Lemeshow test was used to assess the goodness of fit of the model. The variance inflation factor (VIF) was used to detect the presence of multiple collinearity among the model predictors; collinearity was not detected among variables. The Mann-Whitney U test was used to assess the distribution of coagulation profiles among survivors and non-survivors. All tests were performed as two-tailed and statistical significance levels set at a *P* value < .05 in all applied analyses using Stata (StataCorp version 16, College station, Texas).

## RESULTS

A total of 172 patients tested positive for SARS-CoV-2 between March 12^th^ and May 8^th^, 2020. Men were slightly overrepresented as compared to women (96 [55.8%] versus 76 [44.2%], respectively). The median age was 53 years (range: 33.5 – 68 years). A significant majority of patients were under the age of 61 years (113 [65.7%]). Hospitalization was required for 121 patients (70.3%); 51 patients (29.7%) were discharged home and instructed to self-isolate. Among all 121 patients who were hospitalized, 28 (23.1%) were treated in the intensive care unit and 73 (60.3%) were successfully discharged. A total of 18 patients (10.5%) died.

Review of the distribution of cases based on ethnicity revealed that 87 individuals (50.1%) identified as Hispanic and 85 (49.4%) identified as non-Hispanic. Notably, Hispanics account for only 25.7% of the population of Northern Nevada [7]. Hispanic individuals were significantly younger than their non-Hispanic counterparts, with a mean age of 55.8 years versus 46.0 years, respectively. Furthermore, 68 (78.2%) Hispanic patients were under the age of 61 years whereas only 45 (52.9%) of non-Hispanic patients were under the age of 61 years (Table 1).

**Table 1:** Baseline Characteristics and Outcomes According to Ethnicity.

|  | Patients, No (%) |  | p-value |
| --- | --- | --- | --- |
|  | Non-Hispanic | Hispanic |  |
| **Age** |  |  |  |
| Less than 61 years | 45(52.94) | 68(78.16) | 0.001 |
| 61 years and above | 40(47.06) | 19(21.84) |  |
| **Sex** |  |  |  |
| Male, n=96 | 46(54.12) | 50(57.47) | 0.759 |
| Female, n=76 | 39(45.88) | 37(42.53) |  |
| **Income Class** |  |  |  |
| Low income | 26(30.59) | 46(52.87) | 0.003 |
| Middle/Upper income | 59(69.41) | 41(47.13) |  |
| **Primary care provider (PCP)** |  |  |  |
| Has a PCP | 52(61.18) | 37(42.53) | 0.015 |
| Does not have PCP | 33(38.82) | 50(57.47) |  |
| **Insurance Type** |  |  |  |
| Medicare | 28(32.94) | 9(10.34) | <0.001 |
| Medicaid | 15(17.65) | 15(17.24) |  |
| Private | 32(37.65) | 32(36.78) |  |
| Uninsured | 7(8.24) | 24(27.59) |  |
| Workman's comp | 3(3.53) | 7(8.05) |  |
| **Comorbidities** |  |  |  |
| Hypertension | 45(52.94) | 25(28.74) | 0.002 |
| Diabetes | 23(27.06) | 26(29.89) | 0.737 |
| Dyslipidemia | 29(34.52) | 16(18.39) | 0.023 |
| Obesity (BMI > 30) | 46(54.12) | 43(49.43) | 0.546 |
| Coronary artery disease | 3(3.53) | 0 | 0.119 |
| Congestive heart failure | 9(10.59) | 2(2.3) | 0.031 |
| Atrial fibrillation | 8(9.41) | 1(1.15) | 0.017 |
| Cerebrovascular accidents | 11(12.94) | 2(2.30) | 0.009 |
| Chronic Kidney Disease | 11(12.94) | 5(5.75) | 0.121 |
| Asthma | 5(5.88) | 3(3.45) | 0.494 |
| COPD | 10(11.76) | 0(0.00) | 0.001 |
| History of deep venous thrombosis | 4(4.71) | 2(2.30) | 0.441 |
| Cancer | 10(11.76) | 2(2.30) | 0.017 |
| Immunosuppression | 12(14.12) | 2(2.30) | 0.005 |
| Obstructive Sleep apnea | 3(3.53) | 3(3.45) | 1 |
| **Outcomes** |  |  |  |
| Hospitalization | 63(74.12) | 58(66.67) | 0.319 |
| ARDS | 19(22.35) | 14(16.09) | 0.336 |
| Sepsis | 20(23.53) | 21(24.14) | 1 |
| Shock | 11(12.94) | 8(9.20) | 0.474 |
| Mechanical Ventilation | 10 (11.76) | 9(10.34) | 0.812 |
| Hepatic injury | 16(18.82) | 13(14.94) | 0.545 |
| Renal injury | 10(11.76) | 7(8.05) | 0.453 |
| Arrhythmia | 6(7.06) | 5(5.75) | 0.765 |
| Coagulopathy | 31(36.47) | 32(36.78) | 1 |
| Bleeding | 1(1.18) | 3(3.45) | 0.621 |
| Death | 13(15.29) | 5(5.75) | 0.048 |

Three comorbidities were found to significantly increase the risk of mortality among patients with COVID-19: obesity, hypertension, and chronic obstructive pulmonary disease (Table 2). Hypertension and chronic obstructive pulmonary disease were significantly more common among non-Hispanics as compared to Hispanics (45 [52.9%] versus 25 [28.7%] and 10 [11.8%] versus 0 [0%], respectively). Other comorbidities that were more common among the non-Hispanic population include dyslipidemia, obesity, coronary artery disease, congestive heart failure, atrial fibrillation, cerebrovascular accidents, chronic kidney disease, asthma, cancer, and immunosuppression. However, the total sample size was not large enough to calculate the effect of these comorbidities on mortality.

**Table 2:** Multivariate Logistic Regression Analysis of Mortality.

| **Predictor Variable** | **Odds Ratio** | **95% Confidence Interval** | **p-value** |
| --- | --- | --- | --- |
| Female | 0.58 | 0.09 - 3.66 | 0.57 |
| Age >61 years | 5.63 | 0.41 - 77.51 | 0.2 |
| Hispanic | 0.29 | 0.03 - 3.14 | 0.31 |
| Diabetes | 1.93 | 0.37 - 10.11 | 0.44 |
| Hypertension | 17.02 | 1.30 - 223.39 | 0.03 |
| Obesity (BMI> 30) | 10.55 | 1.07 - 104.45 | 0.04 |
| Chronic kidney disease | 1.61 | 0.21 - 12.40 | 0.65 |
| COPD | 15.45 | 1.33 - 178.98 | 0.03 |
| ICU stay | 159.14 | 10.45 -2423.10 | <0.001 |

Median household income was determined based on patient zip code. A total of 72 patients (41.9%) lived in low-income neighborhoods; 100 (58.1%) lived in middle- or upper-income neighborhoods. Hispanic individuals were significantly more likely than non-Hispanics to live in low-income zip codes (46 [52.9%] versus 26 [30.6%], respectively). Hispanics were also significantly more likely to be uninsured and less likely to have a primary care provider as compared to non-Hispanics (24 [27.6%] versus 7 [8.2%] and 37 [42.5%] versus 52 [61.2%], respectively).

Mortality was nearly threefold lower among Hispanics as compared to non-Hispanics (5 [5.8%] versus 13 [15.3%]); the case fatality rate for Hispanics and non-Hispanics was 0.57% and 1.53%, respectively. However, there was no significant difference in the rate of COVID-19-related in-hospital complications – including sepsis, shock, coagulopathy, and acute respiratory distress syndrome – between the two groups. The prothrombin time (PT) and D-dimer levels were significantly higher in non-survivors as compared to survivors (Figure 1); differences in PT and D-dimer were not significantly different between Hispanics and non-Hispanics (p = 0.06).

**Figure 1:**
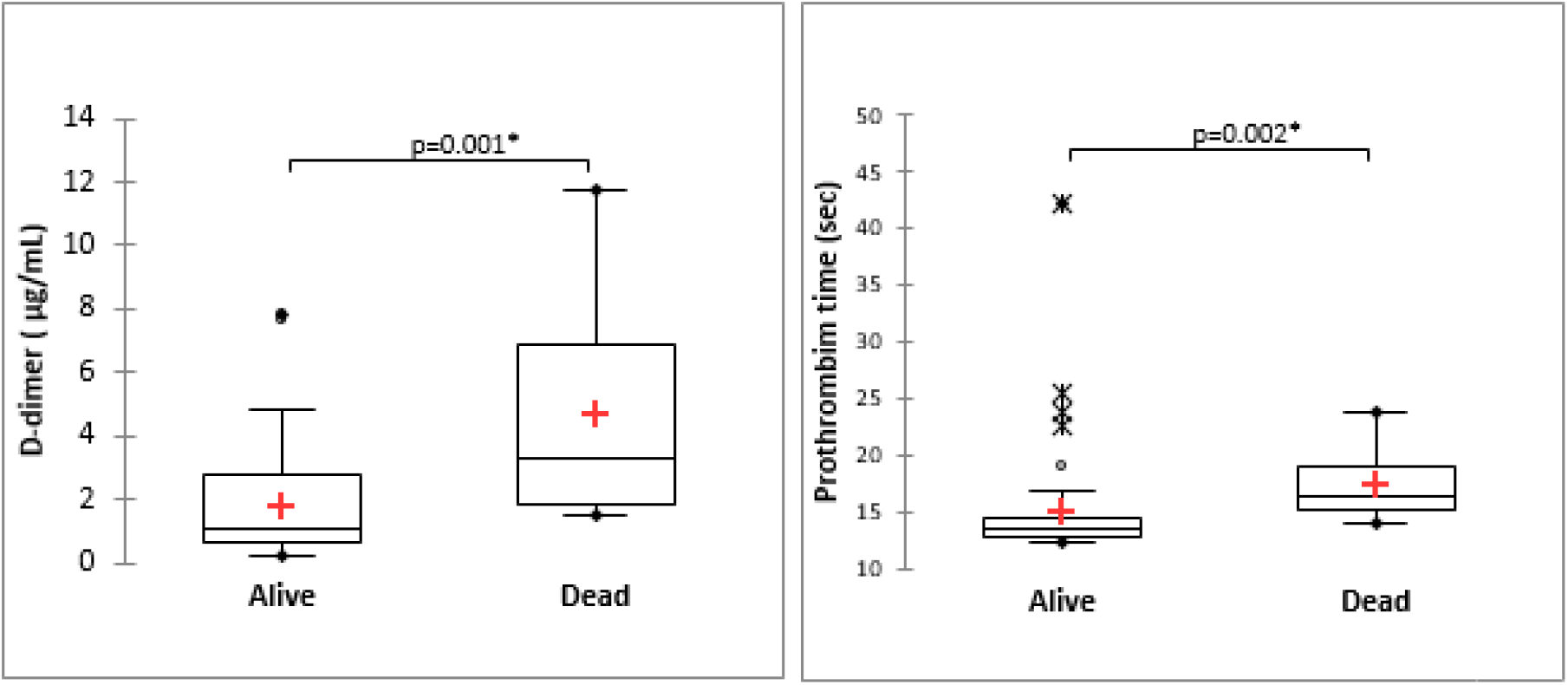
Comparison of coagulopathy profile among survivors and non-survivors. Differences were significant at 5% significance level with the Mann-Whitney U test.

## DISCUSSION

The relationship between ethnicity and COVID-19 represents a public health research priority. Existing data suggest a disproportionate burden of illness and death among minority communities [8]. However, the underlying causes of ethnic disparity in the setting of COVID-19 remain to be definitively established. It has been widely hypothesized that socioeconomic factors – including work circumstances and living conditions – account for the increased incidence and mortality observed among minority communities. Indeed, ethnic minorities are markedly overrepresented in essential industries [9] and often live in multi-generational households [10]; practicing social distancing is therefore unrealistic if not impossible. As Clyde Yancy astutely observed in a recent editorial, “Being able to maintain social distancing… [is an issue] of *privilege*. In certain communities these privileges are simply not accessible [4].”

Our findings demonstrate that Hispanics are disproportionately affected by COVID-19 in Northern Nevada. Hispanic individuals comprise only 25.7% of the population [7] but accounted for over half of the confirmed cases. Hispanic patients were significantly younger than their non-Hispanic counterparts, with a mean age of 46.0 versus 55.8 years, respectively. Interestingly, and perhaps consequently, Hispanics had fewer comorbidites and faced a threefold lower mortality rate than non-Hispanics. We propose that the increased prevalence of COVID-19 among young Hispanics reflects known sociodemographic patterns within the United States. Young Hispanic individuals frequently work in essential industries, such as food service and manufacturing [11], and are therefore unable to practice social distancing. Furthermore, as evidenced by our analysis of patient zip codes, Hispanics were more likely than non-Hispanics to reside in densely-populated, low-income neighborhoods. Increased risk of workplace exposure in the setting of limited social distancing inevitably creates a sociocultural environment favorable to the spread of disease.

It is conceivable that our data underestimate the prevalence of COVID-19 within the Hispanic community. Northern Nevada is home to a small but significant number of undocumented immigrants, and many Hispanic individuals who develop symptoms of COVID-19 may avoid seeking medical care. Indeed, fear of deportation has been established as a major barrier to health care access [12]. In the context of potential under-reporting, the disproportionate burden of disease among Hispanics is particularly striking.

Data on ethic disparity in COVID-19 have demonstrated significantly increased mortality among Hispanics as compared to whites [13-14]; this finding is puzzling in light of the demographics of the United States, where the median age for Hispanics is 27 years versus 37 years for the general population. The age distribution in our study was consistent with national data: Hispanics diagnosed with COVID-19 were approximately one decade younger than their non-Hispanic counterparts. The case fatality rate of COVID-19 increases with age [15], and therefore crude COVID-19-related deaths would be expected to be lower among Hispanics. Furthermore, morbidity and mortality data from the 2009 H1N1 influenza pandemic – a similar global health crisis – indicate that Hispanics were less susceptible to complications than non-Hispanics [16]. In light of the relative youth of the Hispanic populations and mortality patterns observed in the H1N1 influenza pandemic, why has excess COVID-19-related mortality been reported in numerous cities across the United States?

Interestingly, our findings are consistent with those from the neighboring state of California, where the case fatality rate was found to be slightly lower among Hispanics [17]. Studies on social determinants of health have shown that Hispanics face poverty and chronic illness at far greater rates than non-minorities [18] and would therefore be expected to experience excess mortality when compared to age-matched non-Hispanics. Indeed, this was established in a recent report by Gross et al., who concluded that the age-adjusted COVID-19 mortality rate was nearly twice as high in Hispanics as compared to whites [19]. However, in contrast to our findings in Northern Nevada and those reported in California, crude mortality in many cities and states is higher among Hispanics than whites. For instance, in New York City, the crude death rate for Hispanics is 21.3 per 100,000 as compared to 15.7 per 100,000 in non-Hispanic whites and 9.1 per 100,000 in non-Hispanic Asians [14]. Disparity of this magnitude warrants further investigation. We hypothesize that the comparatively low population density and disease burden of some states, such as California and Nevada, allows for greater access to health care than the urban epicenters; this may manifest as improved survival among minorities, who are conceivably less likely to delay treatment in the setting of equitable resource allocation.

It has also been postulated that susceptibility to infection may vary based on ethnic variation in allele distribution of the androgen receptor [20]. Biomolecular studies have demonstrated that the transmembrane protease, serine 2 (TMPRS22), which is regulated by androgen receptors, is required for SARS-CoV-2 infectivity [21]. Furthermore, epidemiologic data – including the present study – have established that COVID-19-related mortality is higher in men as compared to women [22]. It is therefore feasible that Hispanics express a variant of the androgen receptor that transcribes TMPRS22 at a higher rate than non-Hispanics.

Community interventions targeting Hispanic communities have the potential to decrease the incidence of COVID-19 among this disproportionately affected and historically underserved group. Nearly one-fifth of Hispanic Americans speak only Spanish [23]; social distancing messaging should therefore be broadcast in both English and Spanish, particularly in Hispanic-predominant counties. Foreclosures and evictions should be suspended in order to prevent an increase in multi-individual and multi-generational households. Individuals in essential industries should be provided with resources that allow for telecommuting where possible. In an increasingly interconnected world, mitigating ethnic disparity improves the health not only of the Hispanic community, but of all communities nationwide.

In conclusion, our single-center, retrospective observational study revealed that Hispanic individuals are disproportionately affected by COVID-19 in Northern Nevada. Hispanics tended to be younger and have fewer comorbidities than their non-Hispanic counterparts. Consequently, despite social determinants of health that would predict poorer outcomes – including low socioeconomic status, lack of insurance, and absence of a primary care provider – Hispanics experienced a significantly lower case fatality rate than non-Hispanics. The underlying causes of these ethnic disparities warrant further investigation. However, in light of the marked overrepresentation of Hispanics in the COVID-19-positive population of Northern Nevada, targeted public health messaging has the potential to improve outcomes for the Hispanic community as well as the population at large.

## Data Availability

The datasets generated during and/or analyzed during the current study are available from the corresponding author on reasonable request.

